# Obesity Related Glomerulopathy: weighing in the effect of Body Surface Area

**DOI:** 10.1101/2021.03.11.21253278

**Authors:** Bielopolski Dana, Singh Neha, S Bentur Ohad, Renert-Yuval Yael, MacArthur Robert, S Vasquez Kimberly, S Moftah Dena, D Vaughan Roger, G Kost Rhonda, N Tobin Jonathan

## Abstract

**Importance:** Obesity-related glomerulopathy (ORG), part of the cardio-renal spectrum, has an early reversible stage of hyperfiltration. Early identification in the obese adolescent population provides an opportunity to reverse the damage.

**Objective:** Age-appropriate formulae for estimated glomerular filtration rate (eGFR), are standardized to ideal body surface area (BSA) and provide assessment of renal function in mL/min/1.73 m^2^ units, may underestimate prevalence of early ORG. We investigated whether adjusting eGFR to actual BSA more readily identifies early ORG.

**Design:** Cross sectional cohort study. Data were collected between 2011-2015 and analysis was performed between January-November 2020.

**Setting:** Electronic health records clinical data base from 12 academic health centers and community health centers in the New York metropolitan area.

**Participants:** 22,417 women and girls ages 12-21 years for whom data of body measurements and renal function were available.

**Main Outcome and measures:** The hypothesis was generated using previously collected health record data. eGFR was calculated in two ways: BSA-standardized eGFR according to KDIGO recommended formula; and Absolute eGFR adjusted to individual BSA. Hyperfiltration was defined above a threshold of 135mL/min/1.73 m^2^ or 135 mL/min, respectively. The prevalence of hyperfiltration according to each formula was assessed in parallel to 24-hour urine creatinine.

**Results:** 22,417 female adolescents mean age 17 with high prevalence of underrepresented populations (32.6% African American, 12.8% Caucasians and 40.4% Hispanic) were evaluated. Serum creatinine values and hyperfiltration rates according to BSA-standardized eGFR were similar,13.4-15.3%, across Body Mass Index (BMI) groups. Prevalence of hyperfiltration determined by Absolute eGFR differed across groups: Underweight – 2.3%; Normal 6.1%; Overweight – 17.4%; Obese – 31.4%. This trend paralleled the rise in 24-hour urine creatinine across BMI groups.

**Conclusions and relevance:** Absolute eGFR more readily identifies early ORG compared to currently used formulae, which are adjusted to an archaic value of a BSA, not representative of current population BMI measures. The high proportion of underrepresented populations in this study accounts for the higher-than-expected obesity rates and should raise awareness for missed opportunities for screening, early diagnosis, and intervention particularly in young Black adults.

**Key points:** *Question:* Do the currently recommended formulae estimating GFR reliably predict hyperfiltration due to Obesity Related Glomerulopathy (ORG)?

*Findings:* Renal function in relation to BMI was evaluated in a cohort of 22,417 adolescents from the New York metropolitan. Serum creatinine values and BSA-standardized eGFR (mL/min/1.73m^2^) were similar across BMI groups, and as a result, hyperfiltration rates were also similar. However, Absolute eGFR (mL/min) adjusted to individual BSA, created a positive trend across BMI groups similar to urine creatinine.

*Meaning:* Absolute eGFR better reflects the prevalence of hyperfiltration due to Obesity Related Glomerulopathy providing an opportunity for early intervention and damage reversal.

## Introduction

Obesity has reached epidemic proportions in the United States over the past three decades with significant increases in rates of obesity and severe obesity among adolescent females ages 16 to 19 years.^1^,^2^ Obesity is a modifiable risk factor for both chronic kidney disease (CKD) and end-stage renal disease (ESRD).^1^ In children, significant increases in the prevalence of CKD and ESRD have been reported over the last three decades, paralleling the increase in the prevalence of childhood obesity.^1,3^ Obesity-related glomerulopathy (ORG) is a secondary form of focal segmental glomerulosclerosis (FSGS) occurring in obese patients with a BMI >30 kg/m^2^, and results from hemodynamic changes, manifesting as glomerular hyperperfusion and hyperfiltration, due to afferent arteriolar dilatation.^4^ It is well established that in the first stages of ORG, hyperfiltration occurs as a physiological adaptation of the kidney to the increased body mass.^4^ Several studies have reported that obesity-associated hyperfiltration improves after marked weight loss, indicating that hyperfiltration represents a reversible physiologic adaptation.^5^ Although a universal definition of hyperfiltration does not exist, the commonly used threshold is a glomerular filtration rate (GFR) value greater than 135 mL/min/1.73 m^6,7^

### Kidney Disease

Improving Global Outcome (KDIGO) guidelines recommend using age-appropriate serum creatinine-based equations to calculate estimated GFR (eGFR): CKD-Epidemiology Collaboration (CDK-EPI) in the adult patient population and Schwartz in the pediatric patient population. ^8, 9, 10^ However, these equations were empirically developed in populations with renal function impairment (i.e. reduced GFR), and their performance is modest to poor in healthy populations.^10–12^

The gold standard for determining GFR is measurement of exogenous filtration markers, such as inulin, or iothalamate.^13^ However, these are available only as research tools.^14^ Creatinine clearance (CrCl), a readily available measure of GFR, systematically overestimates GFR as a result of net tubular secretion of creatinine.^15^

The eGFR reported by the currently used formulae is standardized to a BSA of 1.73m^2^ rather than the individual’s actual BSA. The value of 1.73 m^2^ reflects the mean BSA of 25-year-old men and women in the United States in 1927^9^ compared to a mean BSA of 1.98 m^2^ in men and women in 2018.^16^ However, the reference value of 1.73m^2^ is maintained for normalization purposes.^9^

We hypothesized that adjusting eGFR to individual BSA, rather than 1.73m^2^, will identify more patients with hyperfiltration in the obese adolescent population, thus making it possible to intervene and reverse the natural course of ORG before attendant kidney damage has long-term, irreversible consequences.

## Methods

### Cohort construction

This study design grew out of an earlier National Institute of Mental Health (NIMH)- funded clinical trial to compare the effectiveness of two different prenatal care strategies among pregnant adolescent women^17^ and a subsequent “big-data” observational study^18^ which examined cardio-metabolic risk factors and birth outcomes among a cohort of adolescent and young women receiving primary care in NYC Health Centers and hospitals. De-identified electronic health record (EHR) data were extracted for female adolescents aged 12-21 years who received health care services from 1/1/2011 to 12/31/2015 in New York City (NYC) in 12 academic health centers and community health centers that are part of Patient Centered Outcomes Research (PCOR)-funded NYC Clinical Data Research Network (NYC-CDRN).^19^

Data extraction and transmission were reviewed and approved, and a waiver of informed consent for analyzing de-identified data was granted by the Institutional Review Boards at Clinical Directors Network (CDN), BRANY (Biomedical Research Alliance of New York), and the Rockefeller University.

Study findings are described in accordance with STROBE guidelines.

### Data-Cleaning steps for biologically plausible limits

The first encounter for each individual where height, weight, blood pressure and serum creatinine were available was included in the cohort. Extreme outlier values that may result from data entry errors rather than true outliers were excluded by setting physiologic limits for systolic blood pressure (60-220 mmHg), diastolic blood pressure (30-150 mmHg), BMI (12-80 kg/m^2^), height (127-200cm), weight (24-240 kg) and serum creatinine (0.3-3 mg/dL).

BSA was calculated using the metric system according to the Du-Bois formula (BSA = 0.007184 * Height^0.725^ * Weight^0.425^).^20^

BMI was calculated by dividing a person’s weight in kilograms by the square of height in meters. BMI values were classified according to the Center for Disease Control and Prevention ^21^ data for BMI-for-age z-score into the following categories: Underweight: BMI ≤5^th^ percentile, Overweight: 95^th^≥BMI≥85^th^ percentile, Obese: BMI ≥95^th^ percentile, all for children and teens of the same age and sex.^21^

### eGFR calculation

The BSA-standardized eGFR was determined by the CKD-EPI and modified Schwartz formulae, for patients aged 18-21 years and 12-18 years, respectively.^10^ Hyperfiltration was defined as BSA-standardized eGFR >135 mL/min/1.73 m^2^.

To eliminate the standardized correction and calculate Absolute eGFR, the BSA-standardized eGFR values were divided by 1.73 and multiplied by individual BSA. This modification was carried out for all patients, regardless of the formula used to calculate eGFR. Hyperfiltration was defined as Absolute eGFR >135 ml/min.^6^

### Statistical Analysis

Statistical analyses were performed using SAS (r 9.4). Nominal variables were expressed as numbers (%). Comparison of proportions between groups was performed using the chi-squared test. Continuous variables were expressed as mean ± SD or median (minimum-maximum). Pearson’s coefficient was used to assess correlation between continuous variables. Continuous variables were compared using ANOVA including Dunnett’s approach for multiple comparisons, where a p<0.05 (two-tailed) was considered statistically significant. Bland-Altman’s analysis was used to assess the relative agreement between BSA-standardized and Absolute eGFR.

## Results

From an original cohort of 123,448 unique patients, following data-cleaning steps to remove biologically implausible values and to verify age and visit dates, 22,417 unique female patients (mean age 17±3 years) with serum creatinine values recorded remained in the final analysis (Supplementary Figure 1). 9,823 (43.8%) of patients were older than 18 years of age, and 12,594 (56.1%) were younger.

Distribution of BMI was sorted into BMI-for-age categories according to CDC^21^ definitions: underweight: 1,085 (4.8%), normal weight: 11,971 (53.4%), overweight: 4,353 (19.4%), and obese: 5,008 (22.3%). The racial distribution of this cohort was: 7,315 (32.6%) African American, and 2,877 (12.8%) Caucasians. 9,068 (40.4%) were Hispanic. African Americans were overrepresented in the obese group compared to Caucasians, (38.2% vs 7.9%, respectively; p<0.001) and underrepresented in the underweight group (27.5% vs 20.6%, respectively; p<0.001) (Table 1).

**Table 1:**
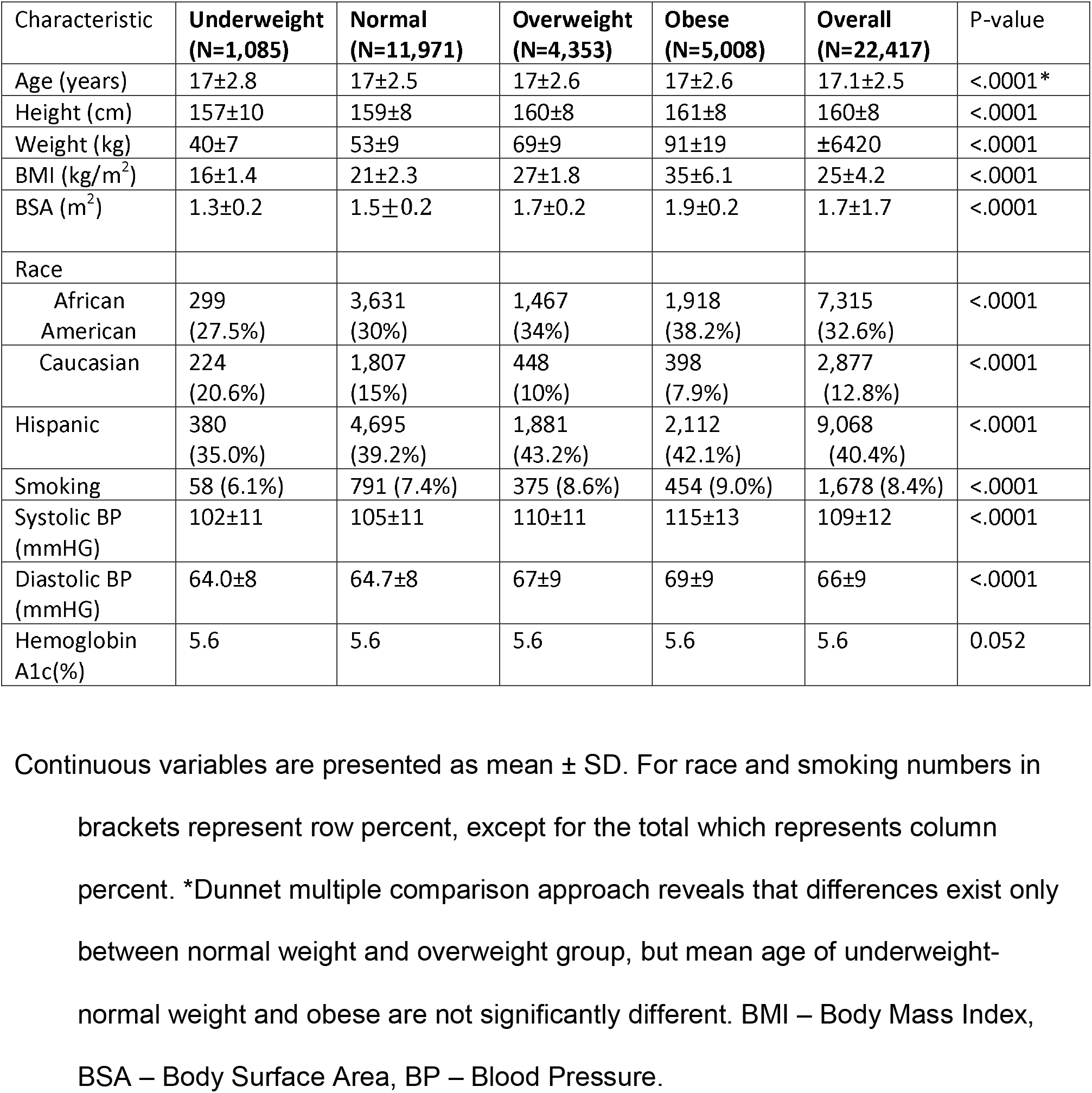
Demographic and vital characteristics of the population.

| Characteristic | **Underweight**<br>**(N=1,085)** | **Normal**<br>**(N=11,971)** | **Overweight**<br>**(N=4,353)** | **Obese**<br>**(N=5,008)** | **Overall**<br>**(N=22,417)** | P-value |
| --- | --- | --- | --- | --- | --- | --- |
| Age (years) | 17±2.8 | 17±2.5 | 17±2.6 | 17±2.6 | 17.1±2.5 | <.0001* |
| Height (cm) | 157±10 | 159±8 | 160±8 | 161±8 | 160±8 | <.0001 |
| Weight (kg) | 40±7 | 53±9 | 69±9 | 91±19 | ±6420 | <.0001 |
| BMI (kg/m<sup>2</sup>) | 16±1.4 | 21±2.3 | 27±1.8 | 35±6.1 | 25±4.2 | <.0001 |
| BSA (m<sup>2</sup>) | 1.3±0.2 | 1.5±0.2 | 1.7±0.2 | 1.9±0.2 | 1.7±1.7 | <.0001 |
| Race |  |  |  |  |  |  |
| African American | 299<br>(27.5%) | 3,631<br>(30%) | 1,467<br>(34%) | 1,918<br>(38.2%) | 7,315<br>(32.6%) | <.0001 |
| Caucasian | 224<br>(20.6%) | 1,807<br>(15%) | 448<br>(10%) | 398<br>(7.9%) | 2,877<br>(12.8%) | <.0001 |
| Hispanic | 380<br>(35.0%) | 4,695<br>(39.2%) | 1,881<br>(43.2%) | 2,112<br>(42.1%) | 9,068<br>(40.4%) | <.0001 |
| Smoking | 58 (6.1%) | 791 (7.4%) | 375 (8.6%) | 454 (9.0%) | 1,678 (8.4%) | <.0001 |
| Systolic BP (mmHG) | 102±11 | 105±11 | 110±11 | 115±13 | 109±12 | <.0001 |
| Diastolic BP (mmHG) | 64.0±8 | 64.7±8 | 67±9 | 69±9 | 66±9 | <.0001 |
| Hemoglobin A1c(%) | 5.6 | 5.6 | 5.6 | 5.6 | 5.6 | 0.052 |

Continuous variables are presented as mean ± SD. For race and smoking numbers in brackets represent row percent, except for the total which represents column percent. \*Dunnet multiple comparison approach reveals that differences exist only between normal weight and overweight group, but mean age of underweight-normal weight and obese are not significantly different. BMI – Body Mass Index, BSA – Body Surface Area, BP – Blood Pressure.

### Renal characteristics

Mean serum creatinine values were 0.73±0.2 mg/dL, 0.75±0.2 mg/dL, 0.74±0.2 mg/dL, and 0.74±0.2 mg/dL (p-value=0.0001) in the underweight, normal, overweight, and obese groups respectively (Table 2).

**Table 2.** Renal characteristics across BMI groups.

| Characteristic | **Underweight**<br>**(N=1,085)** | **Normal**<br>**(N=11,971)** | **Overweight**<br>**(N=4,353)** | **Obese**<br>**(N=5,008)** | **Overall**<br>**(N=22,417)** | P-value |
| --- | --- | --- | --- | --- | --- | --- |
| Serum creatinine (mg/dl) | 0.73±0.2 | 0.75±0.2 | 0.74±0.2 | 0.74±0.2 | 0.74±0.2 | 0.001* |
| Urine creatinine (G/24h) | 0.57±0.3<br>(10) | 0.95±0.4<br>(124) | 1.17±0.4<br>(62) | 1.26±0.5<br>(113) | 1.09±0.4<br>(309) | 0.001** |
| BSA-standardized eGFR (ml/min/1.73m<sup>2</sup>) | 106±30 | 103±27 | 106±27 | 105±26 | 104±27 | <.0001 |
| Hyperfiltration BSA-standardized eGFR | 167<br>(15%) | 1,445<br>(12%) | 630<br>(15%) | 667<br>(13%) | 2,909<br>(13%) | <.0001 |
| Absolute eGFR (ml/min) | 82±26 | 92±26 | 105±29 | 119±33 | 100±31 | <.0001 |
| Hyperfiltration Absolute GFR | 25<br>(2.3%) | 728<br>(6.1%) | 755<br>(17.4%) | 1,568<br>(31.4%) | 3,076<br>(14%) | <.0001 |

Continuous variables are presented as mean ± SD. Absolute numbers are counted individuals with eGFR>135. Numbers in brackets represent column percent.
\*\*trend analysis was significant with p<0.05.
\*Dunnet multiple comparison approach reveals that normal weight and overweight group are statistically indifferent, but mean creatinine of underweight-normal weight and normal weight-obese are significantly different.
eGFR=estimated GFR for ideal body surface area. Absolute GFR=estimated GFR for individuals' body surface area.

Serum creatinine values were similar across BMI groups, but given our large power as a result of the large sample size, BSA-standardized eGFR values were observed to be statistically different yet clinically indistinguishable.^22^ The mean BSA-standardized eGFR in the obese group was 105±26 ml/min/1.73m^2^ vs 106±27ml/min/1.73m^2^ in the overweight group, 103±27 ml/min/1.73m^2^ in the normal weight group, and 106±30 ml/min/1.73m^2^ in the underweight group (Figure 1A, Table 2). Of note, BSA-standardized eGFR values were similar across BMI groups in both the pediatric and adult groups (Table S2, Supplementary Figure 2). In contrast, there was a statistically significant positive trend in mean Absolute eGFR across BMI groups: 82±26 mL/min in the underweight group, 92±26 mL/min in the normal weight group, 105±29 mL/min in the overweight group, and 119±33 mL/min in the obese group (p<0.05; Figure 1B, Table 2).

**Figure 1.**
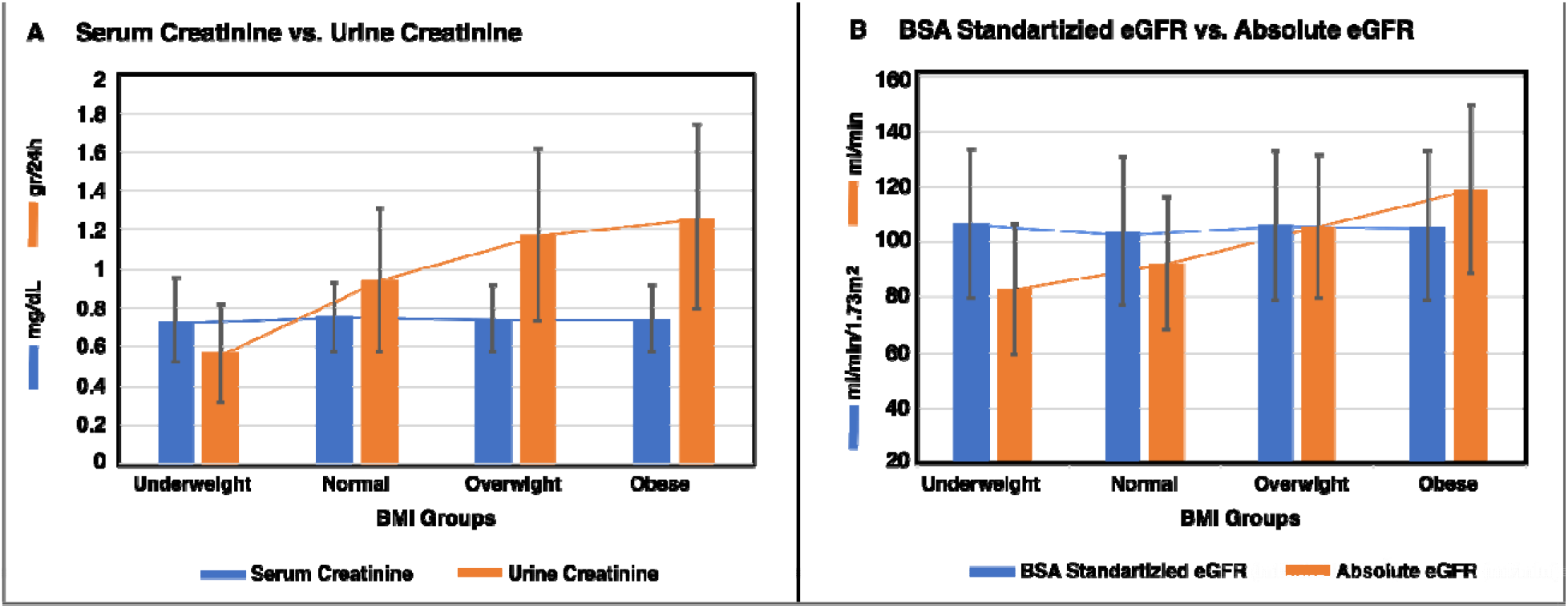
Measured renal characteristics and predicted filtration rate by the eGFR formulae. A) Bars represent mean values of serum creatinine (blue) vs. urine creatinine (orange) for each BMI group. B). Bars represent mean values of predicted filtration according to BSA-standardized eGFR, which is normalized to ideal BSA (blue) vs. Absolute eGFR, which is adjusted to individual BSA (orange). Standard deviations across bars are shown in black lines. Linear lines across bars of the same variable are used to illustrate the trend. p<0.001 for trend of Absolute eGFR and BSA-standardized eGFR across BMI groups, for every unit increase in BMI, Absolute eGFR increases by 2 ml/min compared to 0.3 ml/min/1.73m^2^ for BSA-standardized eGFR. p=0.07 for trend of serum creatinine across BMI groups and p<0.05 for trend of urine creatinine across BMI groups. These are accompanied by an increase in serum creatinine of 0.0002 mg/dL for every unit increase in BMI, whereas for every unit increase in BMI, urine creatinine increases by 0.17 gr/24 hours (equals 170mg/24 hours).

### Hyperfiltration

2,909 patients (12.9%) met the criteria for hyperfiltration according to BSA-standardized eGFR (>135 mL/min/1.73 m^2^): 15.5% from the underweight, 12.0% from the normal weight, 14.5% from the overweight, and 13.3% from the obese groups (p<0.001). Using Absolute eGFR, 3,076 (13.7%) individuals met the criteria for hyperfiltration: 2.3% in the underweight group, 6.1% in the normal weight group, 17.4% in the overweight group and 31.4% in the obese group (Table 2, Figure 2, p<0.001). CrCl data were available for only 74 patients from the entire cohort, hence we used another approach to impute hyperfiltration. Urine creatinine from 24-hour urine collections was available for 309 patients (without urine volume to calculate CrCl). Similar to the trend of increasing Absolute eGFR with increasing BMI, urine creatinine also increased across BMI groups: mean urine creatinine was 0.57±0.25 gr/day in the underweight group, 0.95±0.36 gr/day in the normal weight group, 1.17±0.44 gr/day in the overweight group, and 1.26±0.5 gr/day in the obese group (p<0.05; Figure 1A). Trend analysis confirmed this finding where for every increase in BMI unit urine creatinine increased by 0.02 gr/24 hours (p<0.05; Figure 1A).

**Figure 2.**
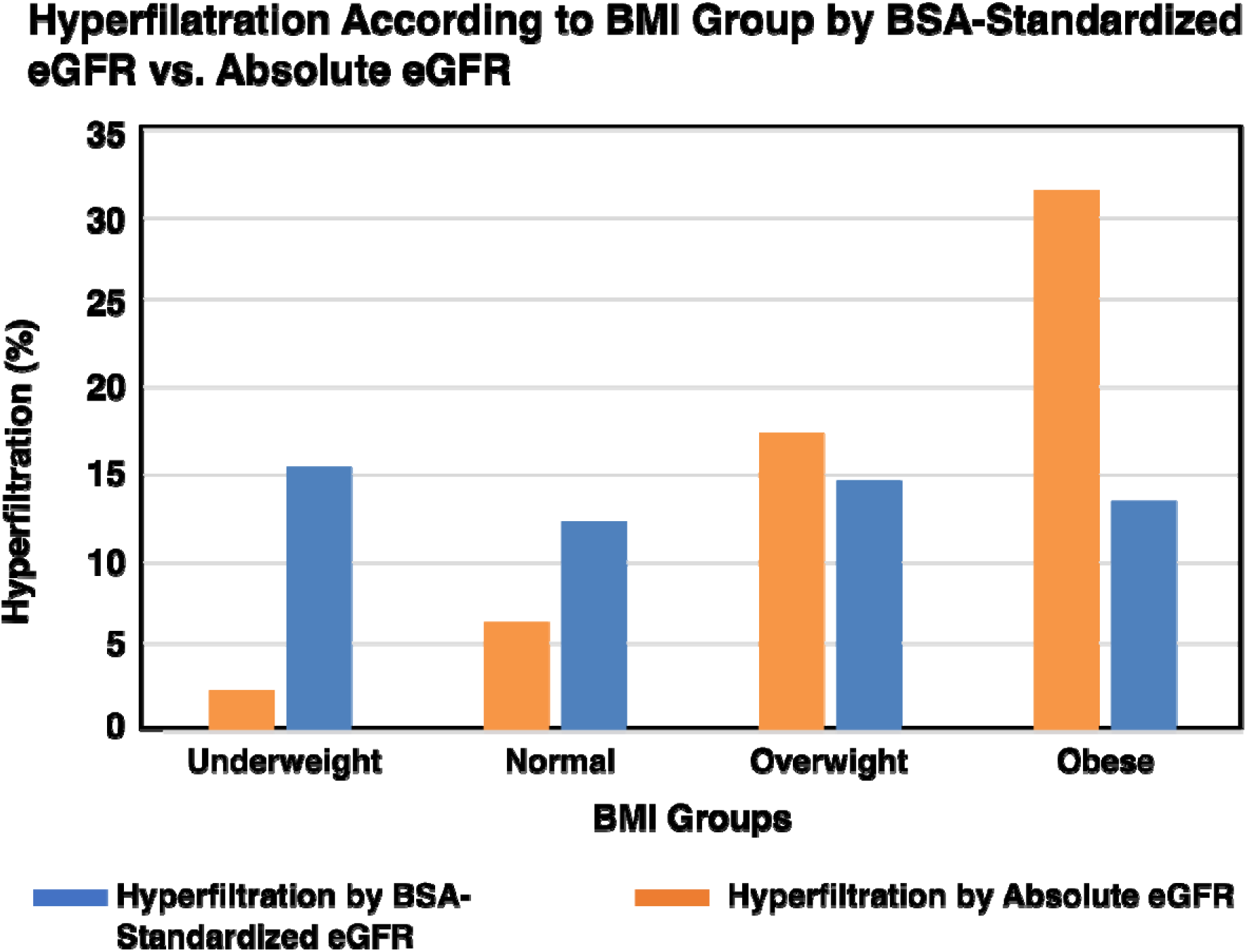
Hyperfiltration prevalence according to BMI group by BSA-standardized eGFR (blue) vs. Absolute eGFR (orange). Hyperfiltration was defined according to GFR threshold of 135 mL/min/17.3m^2^ (BSA-standardized eGFR) or 135 mL/min (Absolute eGFR). (p<0.001 for trend of hyperfiltration according to Absolute eGFR across BMI groups, and p=0.1 for trend of hyperfiltration according to BSA-standardized eGFR across BMI groups)

Bland Altman analyses were performed to test the agreement between BSA-standardized eGFR and Absolute eGFR across the different BMI groups. While there was relatively small negative bias for the overall group (Figure 3e), bias was clearly differential across BMI groups. There was a positive bias for the underweight BMI group, no observed bias for the normal BMI group, an increasing negative bias for the overweight BMI group, and a relatively large negative bias for the obese BMI group (Figure 3a-d).

**Figure 3.**
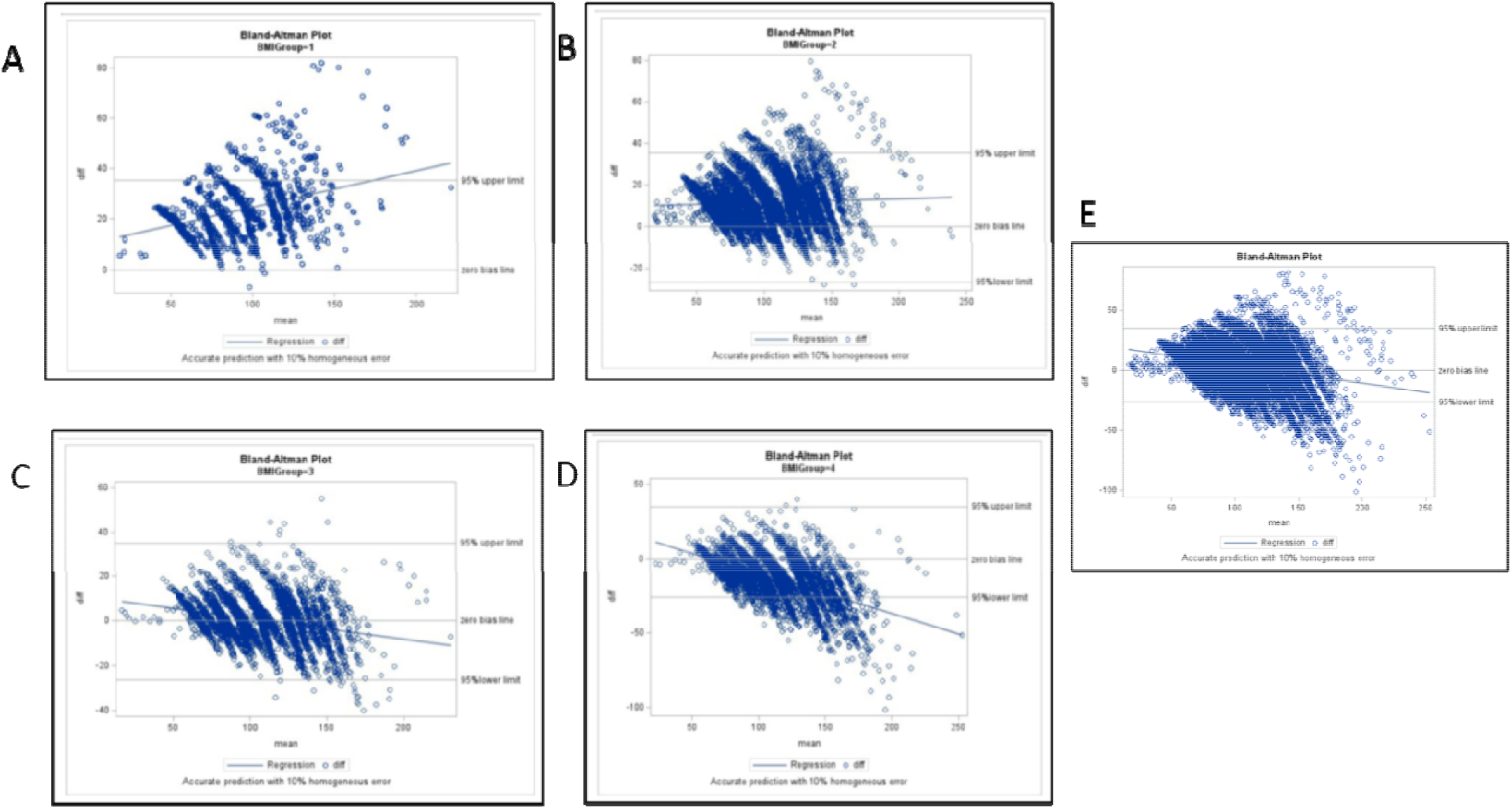
Agreement between BSA-standardized eGFR and Absolute eGFR across BMI groups using the Bland Altman approach. A) Underweight. B) Normal weight. C) Overweight. D) Obese. E) entire cohort.

## Discussion

Accurate calculation of eGFR has a vital role in the diagnosis of kidney disease and CKD management and prognosis. Currently used BSA-standardized eGFR formulae may be adequate in non-obese individuals but might significantly underestimate the true GFR in obese patients,^23^ leading to underdiagnosis of the early stages of ORG, and thus missing an opportunity for intervention. The increasing prevalence of overweight and obesity in children and young adults raises the concern that this metabolic risk gradient probably begins in childhood.

The rationale for using Absolute eGFR is rooted in the pathophysiology of ORG. As body size increases, the number of nephrons remains the same^24^ therefore obesity must result in an increase in single nephron GFR.^25^ Absolute eGFR reflects this phenomenon, whereas ideal BSA (1.73 m^2^)-standardized eGFR obscures it.^26^ As body size increases, the increased single nephron GFR is also burdened by increased sympathetic and renin-angiotensin system activity which leads to an increment of blood pressure, accelerating the progressive deterioration of renal function over time.^27^ We describe a large practice-based cohort of young women and girls ages 12-21 followed in 12 NYC academic and community health centers. While serum creatinine and BSA-standardized eGFR were similar across BMI groups, according to Absolute eGFR, hyperfiltration rates increased as BMI rose in a manner that points to obesity as a possible risk factor for kidney disease. This observation, which may be indicative of a higher prevalence of early stages of ORG, is supported by increased 24-hour urinary creatinine as BMI increased. As BMI rises the kidneys are forced to hyperfiltrate,^28^ however, the CKD-EPI and Schwartz formulae do not reflect this process adequately. The Bland-Altman analysis revealed a lack of agreement between Absolute eGFR vs BSA-standardized eGFR in the obese group and supports the notion of underestimation of eGFR and hyperfiltration according to traditional calculations as we have observed. Standardizing eGFR for BSA had minimal influence in non-obese adults, but can have a major impact and influence decision making in overweight individuals.^29^

Of note, when reviewing data of patients whose renal function were not available (Table S2), we noticed our cohort is more affluent for obese and minority patients, revealing the increased awareness among primary care physicians to these risk factors. Yet the currently available formulae do not support early identification of ORG in this population, and hence this opportunity to improve cardio-renal health is missed.

The course and timeline of ORG in the adolescent has not been thoroughly described. On average, GFR decline is minimal prior to age 35 years, after which the rate of decline accelerates,^30^ highlighting the concept of “renal reserve”. The trajectory of GFR decline over time in relation to BMI is very grim. As BMI increases, kidney function declines, and the decline is more rapid with higher BMI.^31^ Over time the risk of incident end-stage renal disease (ESRD) is also increased in the presence of obesity.^32^ Our study is the first to show the turning point during adolescence, when early intervention, based on Absolute eGFR values, could reverse the initial damage of ORG. Creatinine-based eGFR is strongly influenced by body composition^33^ and our findings suggest that body measures, such as BSA, should be incorporated when eGFR is calculated based on serum creatinine concentrations.^34^

Our study has some limitations. First, we lack parallel data of a gold standard filtration marker for validation of our findings. GFR was not measured directly using inulin or radioisotope methods which are considered the best measures of renal function. Clearly, the use of these exogenous markers to estimate GFR is impractical in clinical practice; however, in patients with normal GFR, CrCl calculated from a 24-hour urine collection provides estimates that are very similar to those obtained with inulin or radioisotopes.^35^ In our cohort, CrCl data were available in only 74 patients, while 24-hour urine creatinine was available in 309 patients without urine volume, which is required for CrCl calculation.

While we observed similar serum creatinine concentrations across BMI groups, 24-hour urine creatinine increased across BMI groups (from underweight to obese), and this implies that the prevalence of hyperfiltration increases as BMI increases. Obesity is associated with higher muscle mass,^15,36^ the main determinant of urinary creatinine.^15^ Measure GFR correlates with urinary creatinine, but formulae used to estimate GFR underestimate it more when urinary creatinine is higher.^15^

Thus, in the presence of normal and similar serum creatinine values across BMI groups, increase in urine creatinine excretion parallel to BMI increase, represents the kidney’s effort to filtrate.

Second, eGFR was calculated using formulae that are recommended by KDIGO, which were designed for different age groups. There are additional formulae, based on serum analytes, such as cystatin c or a combination with creatinine, that are not commonly available in clinical care, thus could not be used in this large cohort, which is based on data from electronic health records.

Another limitation is that according to the BSA-standardized eGFR, 15% of the underweight patients who are not at risk for ORG met the criteria for hyperfiltration. Moreover, this finding was not paralleled by an increase in urinary creatinine in these patients, which questions the reliability of this observation. After adjustment to BSA, Absolute eGFR values in the underweight group correspond to the currently defined CKD2 range according to KDIGO guidelines,^9^ yet data were not extracted in the current data-set to explore renal pathology in this BMI group.

Our study has some clear strengths. Our cohort is large and diverse, including urban population with different racial and ethnic composition compared to the American population. Non-Hispanic Whites make up 61% of the country’s population^37^ compared to less than 15% in our cohort. The high proportion of underrepresented populations in our cohort could account at least partially for the higher-than-expected rates of overweight and obesity, 41% in our cohort compared to 20% the general population, in at ages 12-19.^21^ Obesity at ages 2-19 is more common among Hispanics (25.8%) and non-Hispanic Blacks (22.0%) compared to non-Hispanic Whites (14.1%).^16^ The higher rates of obesity among minorities highlight the urgent unmet clinical need for change in asymptomatic screening and health management among populations affected by health disparities, whose clinical management currently does not adequately address the slippery slope of childhood obesity, when early signs of ORG may already be present. The age range of our cohort, although included under the same definition of adolescence, mandated the use of two different formulae to calculate eGFR. However, the relation we describe, between BSA-standardized eGFR, Absolute eGFR and hyperfiltration rates across BMI groups in the entire cohort exists also separately for the two different age groups and related formula.

Of note, the Schwartz formula does not include a race correction factor, but this factor’s relevance has been questioned in recent publications.^38,39^ Considering the absence of this factor, the high prevalence of overweight and obesity among minority youth and the risk to kidney health needs to be recognized.

In sum, the hypothesis behind this investigation is that eGFR equations may perform differently in those who are obese and overweight compared to those with a normal weight. Currently recommended equations for estimating GFR were empirically developed in populations with reduced GFR, and their performance is modest to poor in healthy populations or during early ORG, when hyperfiltration may predominate.

While the United States Preventive Services Task Force stated in its last report that there are insufficient evidence to recommend routine screening for kidney disease in asymptomatic adults,^40^ early identification of individuals with increased cardio-renal risk could provide opportunities for behavioral, lifestyle, ad pharmacologic interventions to improve long term outcomes.

The most severe form of ORG, associated with significant glomerulosclerosis, has a poor prognosis, where up to 30% of affected individuals reach ESRD 2-6 years after development of glomerulosclerosis.^5^ Lifestyle modification and bariatric surgery can reverse hyperfiltration, reducing GFR for patients with eGFR >90 mL/min following the intervention.^41,42 43^

While the increase in the prevalence of hyperfiltration in the obese group is expected given the mathematical adjustment that was performed, our data show that this seemingly simple adjustment may better represent the true distribution of hyperfiltration in the population and prevalence of ORG in this patient group.

Using Absolute eGFR in clinical practice and research may improve the ability to identify, intervene and reverse early ORG. Application of this tool may address missed opportunities for screening, early diagnosis, and intervention in obese adults, including the Black population, among whom renal disease is over represented, and obesity rates are increasing. Successful intervention has the potential to improve quality of life and prevent subsequent comorbidities, along with care costs reduction for a substantial part of the population at risk, where there is a high representation of underserved minorities.

## Data Availability

Data have been provided by the INSIGHT Clinical Research Network and can be made available upon request. The INSIGHT Governance Board will review each request.

## Acknowledgment

We thank Dr. Mark Weiner from Weill Cornell School of Medicine for persistence in access and data quality. We thank Caroline Jiang from the Rockefeller University for supervising and mentoring data analysis.

## Abbreviations used

CKD: Chronic kidney disease
ESRD: End-stage renal disease
ORG: Obesity-related Glomerulopathy
eGFR: estimated Glomerular Filtration Rate
BSA: Body Surface Area
BMI: Body Mass Index
CDK-EPI: CKD-Epidemiology Collaboration
KDIGO: Kidney Disease Improving Global Outcome
CrCl: Creatinine Clearance
Absolute eGFR: actual BSA based eGFR

## Supplementary materials

**Supplementary Figure 1.**
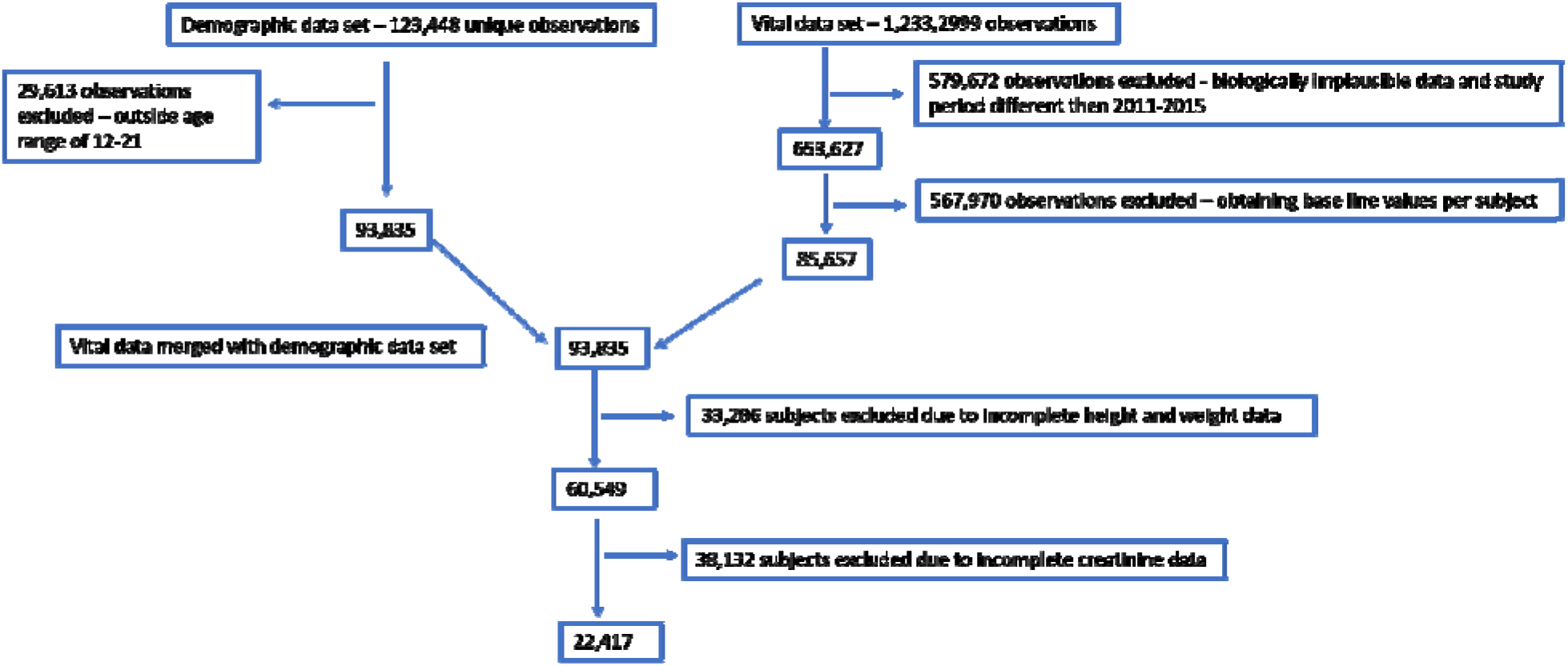
STROBE diagram illustrating cohort construction and data cleaning steps.

**Supplementary Figure 2.**
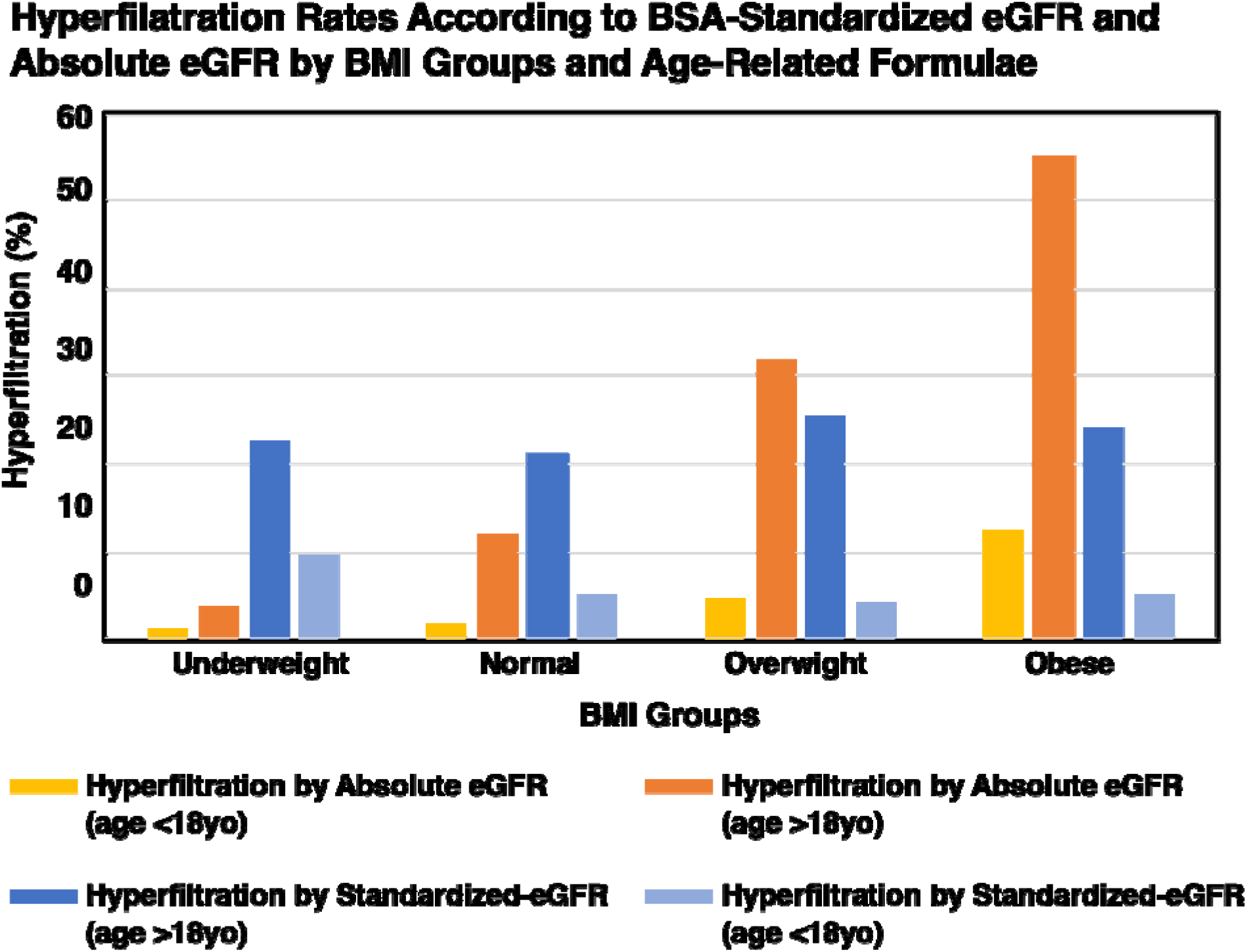
Bars represent rates (%) of hyperfiltration according to age-related formula and adjustment to BSA. (age <18 yo) -age below 18 years where Schwartz formula was used, (age >18 yo) age above 18 years old where CKD-EPI formula was used. (p=0.002 for trend of hyperfiltration rates according to BSA standardized eGFR for patients above age 18, p=0.01 for trend of hyperfiltration rates according to BSA standardized eGFR for patients below age 18, p<0.001 for trend of hyperfiltration rates according to absolute eGFR for patients below age 18, p<0.001 for trend of hyperfiltration rates according to absolute eGFR for patients above age 18)

**Table S1.**
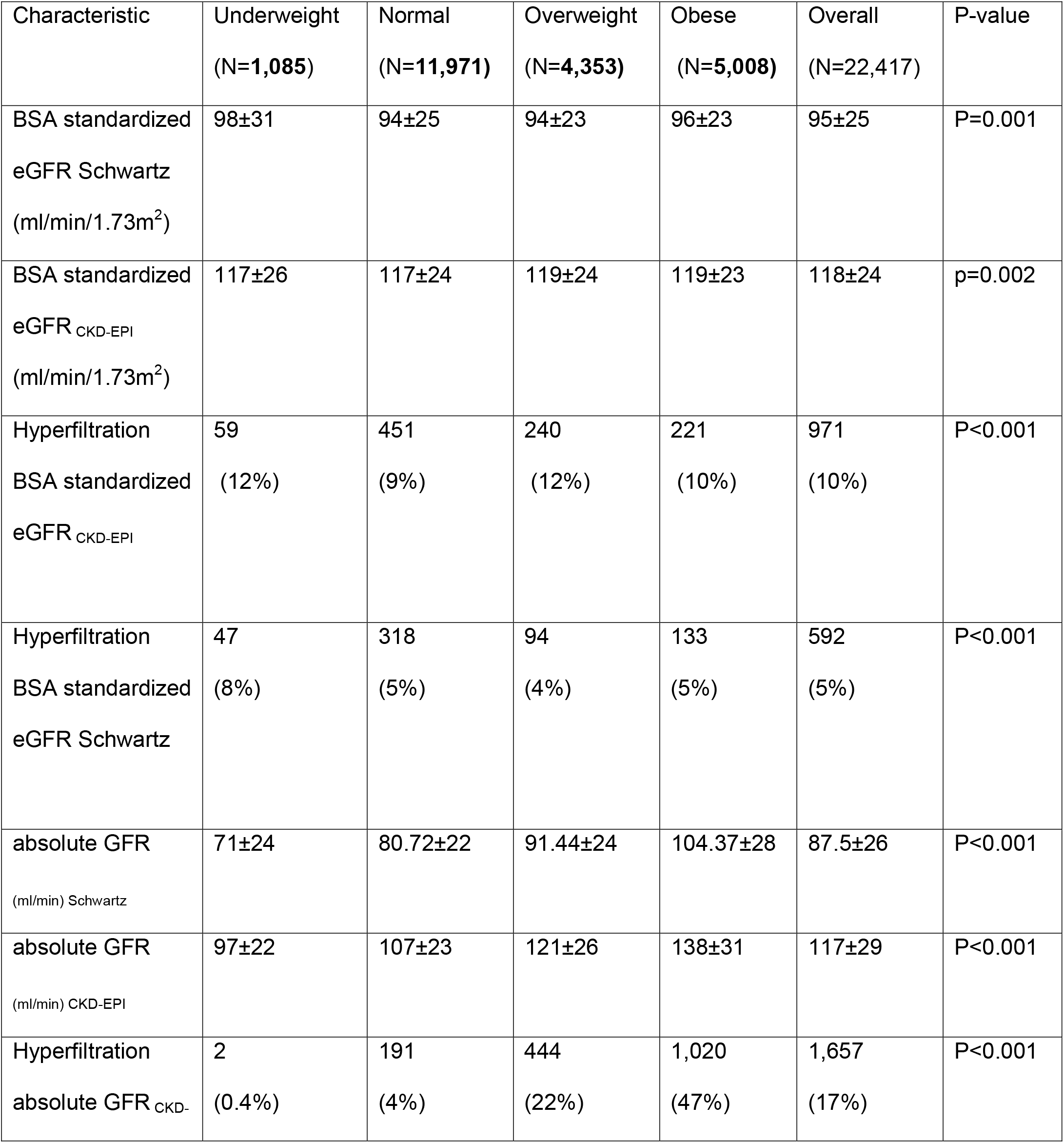
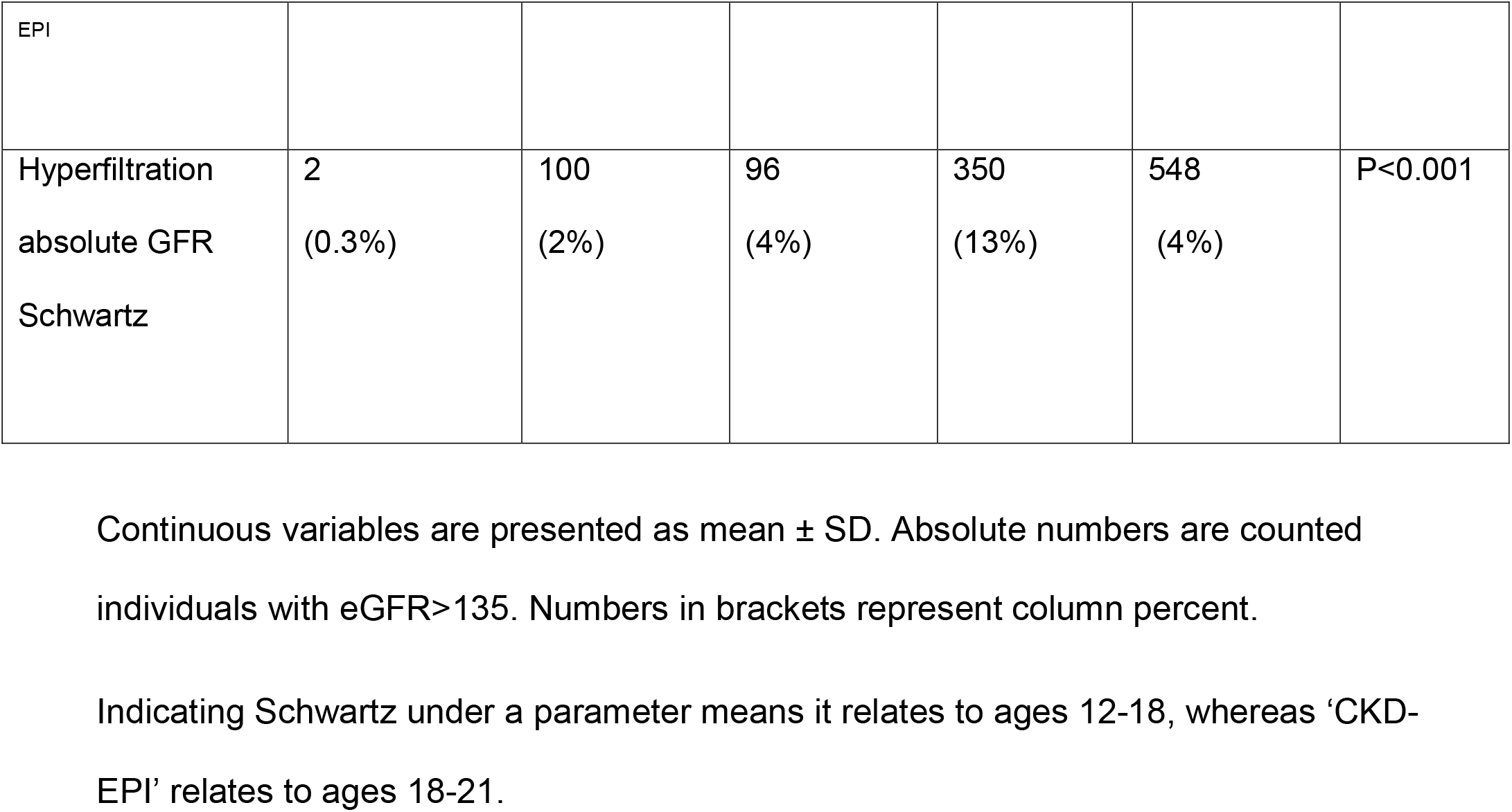
Renal characteristics across BMI groups according to the age specific formulae.

**Table S2.**
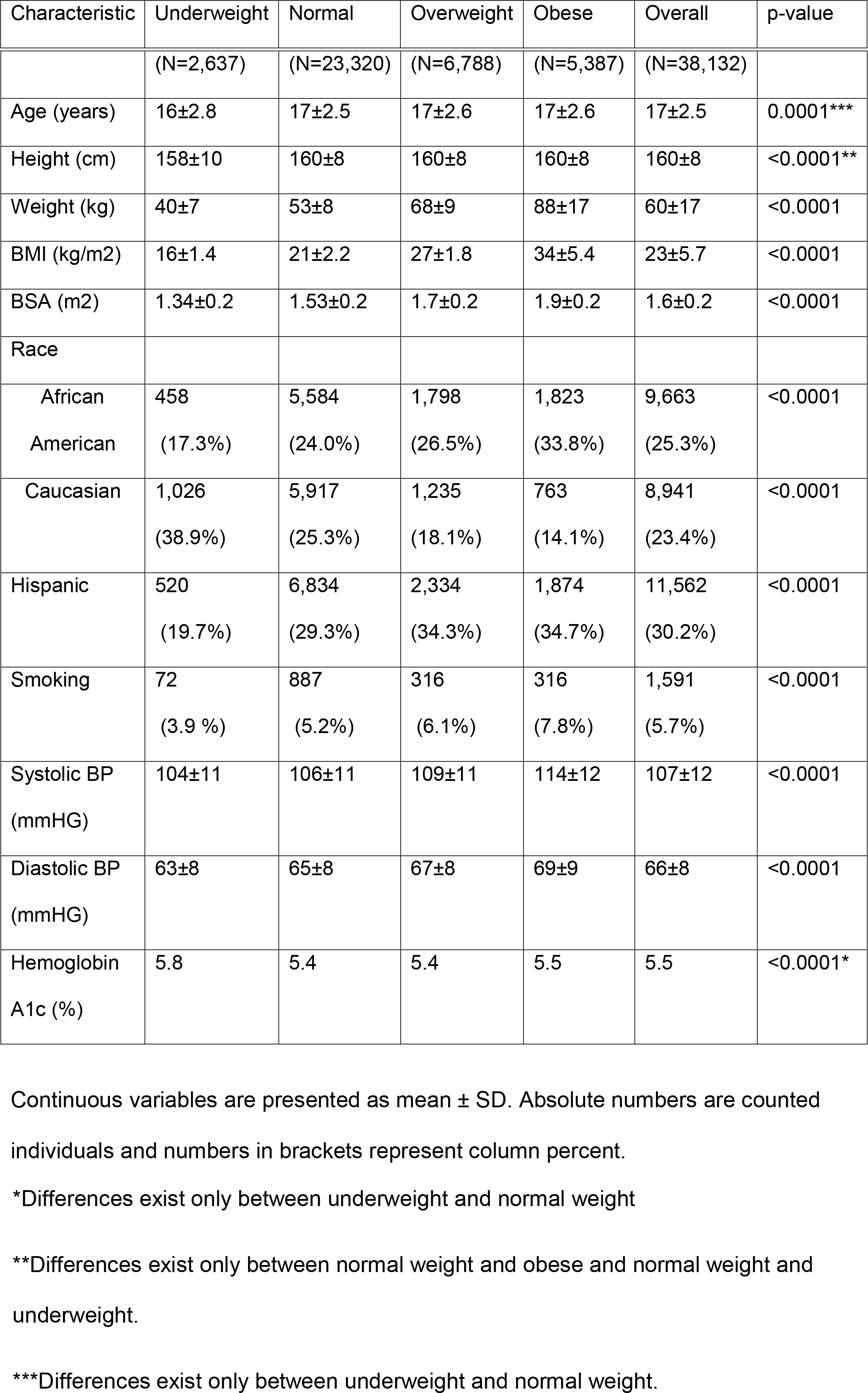
Demographic characterization of patients without documented renal function.

## Table of contents

